# Post-discharge Acute Care and Outcomes in the Era of Readmission Reduction: A National Retrospective Cohort Study of Medicare Beneficiaries in the United States

**DOI:** 10.1101/19002162

**Authors:** Rohan Khera, Yongfei Wang, Susannah M. Bernheim, Zhenqiu Lin, Harlan M. Krumholz

**Affiliations:** Division of Cardiology, University of Texas Southwestern Medical Center, 5323 Harry Hines Blvd, Suite E5.730D, Dallas, Texas, USA 75219.; Department of Internal Medicine, Yale School of Medicine; and; Senior Statistician, Center for Outcomes Research and Evaluation, Yale-New Haven Hospital, 1 Church St., Suite 200, New Haven, Connecticut, USA 06510; Department of Internal Medicine, Yale School of Medicine; and Director, Quality Measurement Programs, Center for Outcomes Research and Evaluation, Yale-New Haven Hospital, 1 Church St., Suite 200, New Haven, Connecticut, USA 06510; Section of Cardiovascular Medicine, Department of Internal Medicine, Yale School of Medicine; and; Senior Statistician, Center for Outcomes Research and Evaluation, Yale-New Haven Hospital, 1 Church St., Suite 200, New Haven, Connecticut, USA 06510; Department of Internal Medicine, Yale School of Medicine; Department of Health Policy and Management, Yale School of Public Health, and; Director, Center for Outcomes Research and Evaluation, Yale-New Haven Hospital, 1 Church St., Suite 200, New Haven, Connecticut, USA 06510

**Author notes:** Correspondence to: Dr. Rohan Khera –; @rohan_khera.

## Abstract

**Background:** With incentives to reduce readmission rates in the United States, there are concerns that patients who need hospitalization after a recent hospital discharge may be denied access, which would increase their risk of mortality.

**Objective:** We determined whether patients with hospitalizations for conditions covered by national readmission programs who received care in emergency department (ED) or observation units but were not hospitalized within 30 days had an increased risk of death. We also evaluated temporal trends in post-discharge acute care utilization in inpatient units, emergency department (ED) and observation units for these patients.

**Design, Setting, and Participants:** In this observational study of national Medicare claims data for 2008-2016, we identified patients ≥65 years hospitalized with heart failure (HF), acute myocardial infarction (AMI), or pneumonia, conditions included in the HRRP.

**Main Outcomes and Measures:** Post-discharge 30-day mortality according to patients’ 30-day acute care utilization. Acute care utilization in inpatient and observation units, and the ED during the 30-day and 31-90-day post-discharge period.

**Results:** There were 3,772,924 hospitalizations for HF, 1,570,113 for AMI, and 3,131,162 for pneumonia. The overall post-discharge 30-day mortality was 8.7% for HF, 7.3% for AMI, and 8.4% for pneumonia. Risk-adjusted mortality increased annually by 0.05% (95% CI, 0.02% to 0.08%) for HF, decreased by 0.06% (95% CI, −0.09% to −0.04%) for AMI, and did not significantly change for pneumonia. Specifically, mortality increased for HF patients who did not utilize any post-discharge acute care, increasing at a rate of 0.08% per year (95% CI, 0.05% to 0.12%), exceeding the overall absolute annual increase in post-discharge mortality in heart failure, without an increase in mortality in observation units or the ED. Concurrent with a reduction in 30-day readmission rates, 30-day observation stays and visits to the ED increased across all 3 conditions during and beyond the post-discharge 30-day period. There was no significant change in overall 30-day post-acute care utilization.

**Conclusions:** The only condition with an increasing mortality through the study period was HF; the increase preceded the policy and was not present among those received ED or observation unit care without hospitalization. Overall, during this period, there was not a significant change in the overall 30-day post-discharge acute care utilization.

**What is already known on this topic:**

- Among Medicare beneficiaries hospitalized for heart failure, mortality in the 30-day post-discharge period has been increasing over the past several years. However, the relationship between post-discharge acute care and mortality in the early post-discharge period remains poorly understood.
- Observation units and the emergency departments (ED) have increasingly been used as avenues for patient care in the United States. However, the utilization of these services in the early post-discharge period for conditions targeted in the Hospital Readmissions Reduction Program (HRRP), and the outcomes of patients in these settings, is required to appropriately evaluate the effects of the program.

**What this study adds:**

- Among conditions targeted in the HRRP, patients with heart failure, but not those with acute myocardial infarction or pneumonia, experienced increase in post-discharge 30-day mortality. This increase preceded the announcement of the program and was concentrated among individuals who sought no post-discharge acute care, nearly half of whom had been discharged to hospice.
- Despite increasing utilization of observation units and the ED in the post-discharge period, care in these settings was not associated with increased mortality risk.

## BACKGROUND

The announcement and implementation of the United States Hospital Readmissions Reduction Program (HRRP) were associated with a reduction in readmissions within 30 days of discharge for heart failure, acute myocardial infarction (AMI), and pneumonia,(1-4) as demonstrated by a decrease in the overall national rate of readmissions. There are concerns that pressures to reduce readmissions have led to the evolution of care patterns that may have adverse consequences through reducing access to care in appropriate settings.(5-7) Therefore, it is essential to determine whether patients who are seen in acute care settings, but not hospitalized, experienced an increased risk of mortality.

There is also a question about the association of the HRRP with the overall use of acute services in the 30-day window. The reduction in readmissions could have occurred in the setting of changes that improved the recovery of patients and reduced the occurrence of clinical events requiring acute care in the early post-discharge period. Alternatively, the decrease in readmissions might not have reduced the need for acute care but instead influenced clinicians to direct patients to the emergency department (ED) or observation units instead of pursuing hospital admission. The care of patients in the ED and observation units is appropriate only if they can be treated adequately in these settings without pursuing admission. An assessment of how outcomes of patients in these care settings has evolved is essential to address concerns regarding the appropriateness of changes in patterns of acute care in the post-discharge period during the application of incentives to reduce readmission.

Accordingly, we determined if the patients who sought acute care in the 30 days after discharge for heart failure, AMI, and pneumonia, the conditions the US Congress included in the HRRP(3, 8, 9), but did not undergo hospitalization, had an increased risk of mortality. In particular, we investigated if people who had a post-discharge emergency department visit that did not lead to a hospitalization had an increase in mortality, addressing whether the incentive programs reduced access to the hospital and caused harm. We examined the characteristics of patients, and the change over time, by their type of acute care in the 30 days after hospitalization.

We also evaluated the temporal trends in the utilization of post-discharge acute care, including readmissions, observation stays and ED visits. We determined if declines in readmissions were associated with increases in other types of acute care. We tested if there were differences over time in 30-day need for acute care. We also evaluated the period from day 31 through day 90 to determine if there was a difference after the evaluation period of HRRP, to see if any practice change was limited to the period covered by the incentives.

## METHODS

### Data Source

We used the Medicare Standard Analytic Files that included 100% inpatient and outpatient claims for the years 2008–2016. The study period spanned the announcement and subsequent implementation of the HRRP in 2010 and 2012, respectively.(8) For each year, we included data for all fee-for-service Medicare beneficiary 65 years of age and older who were hospitalized with a principal discharge diagnosis of heart failure, AMI, or pneumonia. The diagnoses were based on *International Classification of Diseases (ICD)* diagnosis codes that are included in the CMS measures for these conditions and are publicly available.(10-13) Notably, the measure for pneumonia changed in 2014, but to ensure consistency, the study population for pneumonia was defined across the study period using the codes before the measure changed.(12) Further, the coding scheme changed from ICD-9 to ICD-10 in October 2015.(14) Therefore, we used the CMS measure methodology that was rigorously tested for consistency in the patient population for each of 3 conditions across transition.(10)

To align the study cohort with the population of patients under the purview of the HRRP, we excluded individuals who were discharged against medical advice, died during hospitalization, and did not have at least 30-day follow-up after discharge. The cohort was consistent with the specifications of the 30-day readmission metric of the Centers for Medicare & Medicaid Services.(9, 11-13, 15)

### Post-discharge Acute Care Utilization and Mortality

We used revenue center codes specific to claims submitted from observation stays and ED visits to identify these care settings. These included revenue center codes of 0762 and/or Healthcare Common Procedure Coding System code of G0378 for observation stays and revenue center codes of 0450, 0451, 0452, 0459, or 0981 for ED visits based on hospital outpatient claims data. These have been used in a National Quality Forum-endorsed outcome measure focused on post-discharge acute care.(16) We used the Medicare denominator files to identify patients who died from any cause and determined the temporal relationship of their date of death to the day of hospital discharge.(17) We defined 30-day post-discharge mortality as the proportion of individuals discharged alive after hospital discharge who died within 30-days of discharge.

### Post-discharge Care-based Patient Groups

We examined post-discharge mortality in 4 mutually exclusive patient groups that were defined based on the clinical setting of post-discharge care availed by patients in the post-discharge period. These included patients that (1) were readmitted to the hospital within 30 days of discharge for any cause, (2) were not readmitted but had an observation stay in the 30-day post-discharge period for any cause, (3) were neither readmitted nor had an observation stay, but visited the ED for any reason, and (4) had no post-discharge 30-day acute care either as a hospitalization, observation stay, or ED visit.

### Risk-Adjusted Rates of Outcomes

To account for changes in patient characteristics, we constructed logistic regression models with utilization of care and mortality endpoints across care settings as dependent variables and covariates included in the risk-adjustment models in the CMS measures for each condition as independent variables (eTables 1-3). The calendar-year and monthly risk-adjusted rates of utilization and mortality are calculated as the ratio of the observed rate during the period and the expected rate in the period, multiplied by the unadjusted rate over the study period.(9) To ensure consistency in covariates across ICD-9 and ICD-10 coding formats, as a part of the CMS measure design process, combinations of codes within each of the coding schemes were mapped to Condition Category (CC) codes for the covariates included in the model. The ICD-9 and ICD-10 codes included in the CC codes, and data assessments showing consistency of CC codes across the transition, are publicly available.(10)

**Table 1:** Characteristics of patients with heart failure based on survival and post-discharge acute care use. Numbers represent proportion of patients discharged alive with heart failure who either survived to post-discharge day 30 or died during this period. The characteristics of the latter group are presented among groups based on their post-discharge acute-care use.

|  | Alive at Day 30 | Deceased at Day 30 |  |  |  |  |
| --- | --- | --- | --- | --- | --- | --- |
|  |  | Overall | Readmission | Observation stay | ED visit | No readmission/ observation/ED |
| Number of patients | 3,445,780 | 327,144 | 112,673 | 2972 | 23,378 | 188,121 |
| **Demographics** |  |  |  |  |  |  |
| Age, mean (SD), years | 80.5 (8.2) | 83.9 (8.1) | 82.6 (8) | 84 (8.1) | 82.3 (8.4) | 84.8 (7.9) |
| Male sex | 46.1 | 48.0 | 48.9 | 49.6 | 52.1 | 46.8 |
| **Health conditions** |  |  |  |  |  |  |
| Cardio-respiratory failure and shock (CC 79) | 26.9 | 34.6 | 35.8 | 35.5 | 35.4 | 33.7 |
| Dementia (CC 49, 50) | 22.0 | 35.2 | 29.9 | 35.3 | 33.1 | 38.6 |
| Hemiplegia, paralysis, functional disability (CC 67 to 69, 100 to 102, 177, 178) | 7.7 | 9.6 | 9.5 | 9.1 | 9.9 | 9.7 |
| Delirium and encephalopathy (CC 48) | 2.6 | 7.8 | 5.3 | 5.4 | 5.8 | 9.5 |
| Metastatic cancer and acute leukemia (CC 7) | 2.0 | 4.7 | 4.4 | 4.8 | 3.6 | 4.9 |
| Chronic skin ulcer (CC 148, 149) | 13.6 | 21.1 | 20.5 | 18.2 | 20.6 | 21.5 |
| Protein-calorie malnutrition (CC 21) | 8.3 | 18.6 | 16.4 | 16.5 | 16.7 | 20.3 |
| **Care patterns** |  |  |  |  |  |  |
| Intensive care unit use | 47.7 | 52.1 | 50.8 | 47.2 | 51.4 | 53.0 |
| Palliative care consultation | 1.0 | 12.1 | 2.0 | 4.9 | 3.6 | 19.4 |
| Major operative procedures | 5.1 | 3.2 | 4.0 | 3.2 | 4.1 | 2.7 |
| **Discharge disposition** |  |  |  |  |  |  |
| Skilled nursing facility | 19.4 | 32.2 | 40.3 | 36.8 | 40.6 | 26.1 |
| Hospice | 1.4 | 26.9 | 1.4 | 6.1 | 5.0 | 45.3 |
| Home health care | 23.9 | 13.9 | 22.6 | 23.4 | 20.5 | 7.7 |
| Long-term acute-care facility | 0.7 | 2.0 | 1.1 | 0.3 | 1.0 | 2.8 |

ED: emergency department, SD: standard deviation

**Table 2:** Characteristics of patients with acute myocardial infarction based on survival and post-discharge acute-care use. Numbers represent proportion of patients discharged alive with myocardial infarction who either survived to post-discharge day 30 or died during this period. The characteristics of the latter group are presented among groups based on their post-discharge acute care use.

|  | Alive at day 30 | Deceased at Day 30 |  |  |  |  |
| --- | --- | --- | --- | --- | --- | --- |
|  |  | Overall | Readmission | Observation stay | ED visit | No readmission/ observation/ED |
| Number of patients | 1,455,442 | 114,671 | 34,772 | 1094 | 9372 | 69,433 |
| **Demographics** |  |  |  |  |  |  |
| Age, mean (SD), years | 78.2 (8.1) | 83.8 (8.3) | 82.5 (8.2) | 84.1 (8.4) | 81.5 (8.5) | 84.7 (8.2) |
| Male sex | 52.6 | 47.8 | 50.2 | 50.8 | 53.3 | 45.7 |
| **Health conditions** |  |  |  |  |  |  |
| Cardio-respiratory failure and shock (CC 79) | 14.5 | 29.7 | 24.0 | 18.6 | 22.5 | 33.7 |
| Dementia (CC 49. 50) | 17.2 | 38.7 | 30.9 | 36.2 | 32.4 | 43.5 |
| Hemiplegia, paralysis, functional disability (CC 67 to 69, 100 to 102, 177, 178) | 5.7 | 10.0 | 9.6 | 8.6 | 9.9 | 10.2 |
| Delirium and encephalopathy (CC 48) | 3.6 | 9.6 | 7.1 | 7.9 | 6.3 | 11.3 |
| Metastatic cancer and acute leukemia (CC 7) | 1.8 | 5.9 | 5.2 | 5.8 | 4.2 | 6.5 |
| Chronic skin ulcer (CC 148, 149) | 7.2 | 14.5 | 14.3 | 13.7 | 14.3 | 14.7 |
| Protein-calorie malnutrition (CC 21) | 5.1 | 15.4 | 13.1 | 13.8 | 12.4 | 17.0 |
| **Care patterns** |  |  |  |  |  |  |
| Intensive care unit use | 73.2 | 69.4 | 71.7 | 62.7 | 72.3 | 68.0 |
| Palliative care consultation | 0.8 | 13.7 | 2.1 | 6.1 | 3.5 | 21.0 |
| Major operative procedures | 50.8 | 18.2 | 23.8 | 17.6 | 27.0 | 14.3 |
| **Discharge disposition** |  |  |  |  |  |  |
| Skilled nursing facility | 16.6 | 30.7 | 41.0 | 33.7 | 38.4 | 24.4 |
| Hospice | 1.0 | 30.5 | 1.5 | 5.6 | 3.7 | 49.1 |
| Home health care | 16.2 | 10.4 | 18.2 | 20.7 | 16.4 | 5.5 |
| Long-term acute-care facility | 0.8 | 2.8 | 1.9 | 0.7 | 1.2 | 3.6 |

ED: emergency department, SD: standard deviation

**Table 3:**
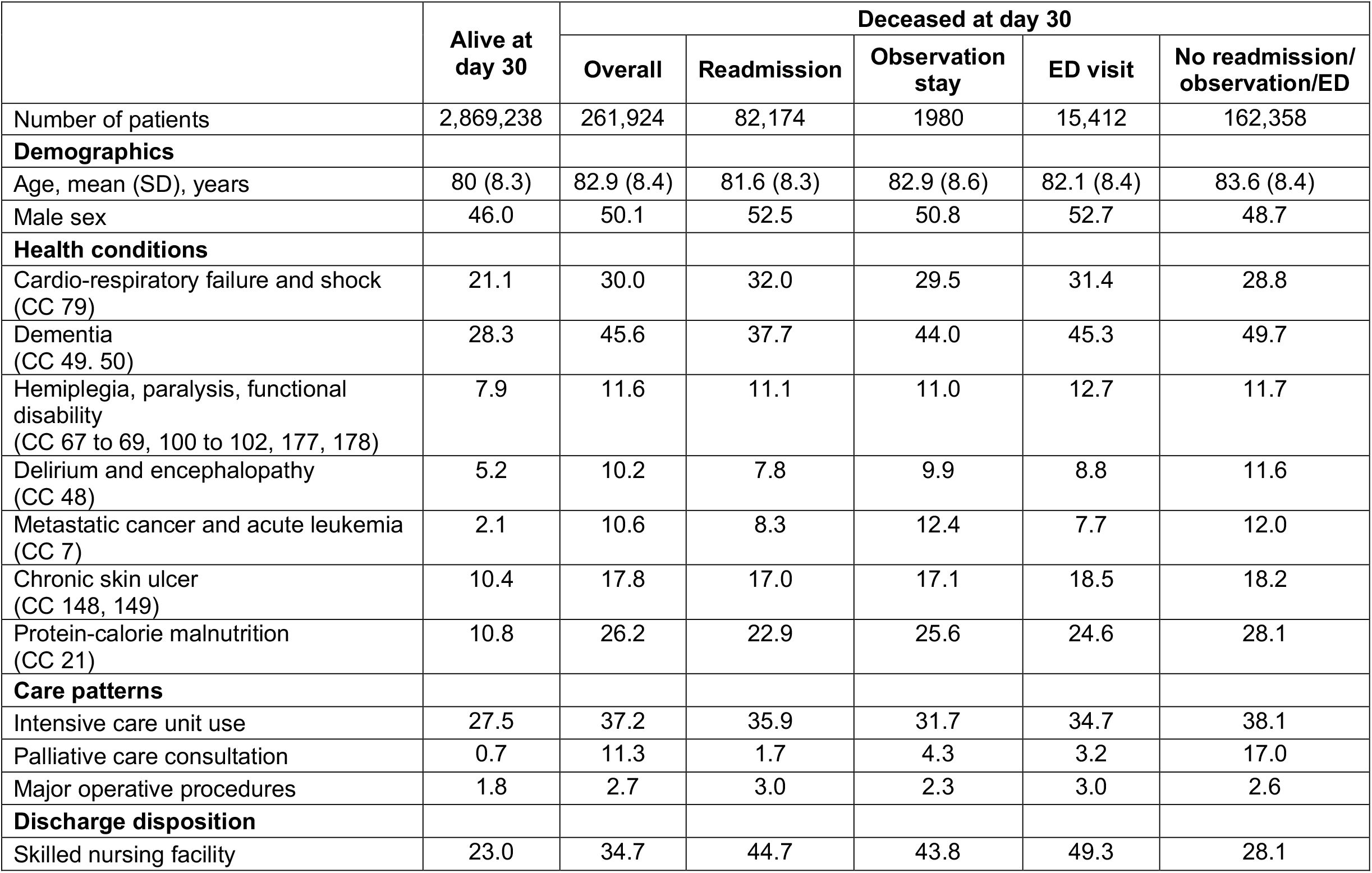
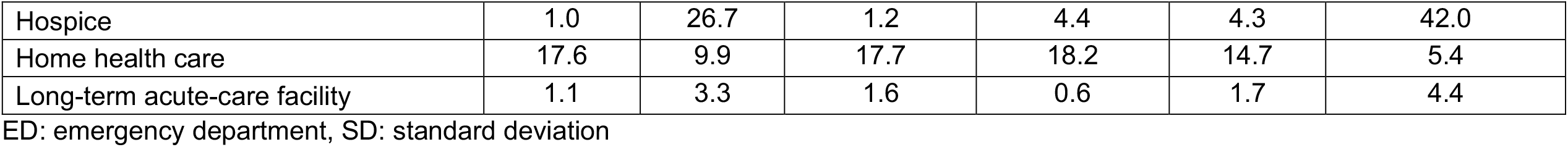
Characteristics of patients with pneumonia based on survival and post-discharge acute-care use. Numbers represent proportion of patients discharged alive with pneumonia who either survived to post-discharge day 30 or died during this period. The characteristics of the latter group are presented among groups based on their post-discharge acute-care use.

There have been changes in the number of codes on inpatient claims over time.(18, 19) We pursued additional analyses to evaluate whether differences in covariate selection strategies substantively altered the results. First, we used inpatient and outpatient claims over the preceding 12-month period to assess covariates. We found that restricting the additional inpatient covariates available from the expansion of codes to the same number available previously did not substantively alter the risk-adjustment models..(20) Nevertheless, to specifically address how the strategy to identify covariates affects risk-adjusted assessments in the current analysis, we created a secondary cohort of hospitalizations with complete information on the order of inpatient claims (95% of the primary cohort), and used two covariate identification strategies, where we combined outpatient claims over the preceding 12 months before the index hospitalization with (a) a fixed number of diagnoses from the claims across study period (9 secondary diagnosis codes, 6 procedure codes), or (b) all inpatient codes (i.e. up to 25 diagnosis and procedure codes after expansion of inpatient code slots).

### Statistical Analysis

First, we evaluated calendar-year and monthly rates of post-discharge care within the 30-day post-discharge period as readmissions, observation stays, and ED visits between 2008 and 2016 for each of the 3 HRRP conditions. Next, for each of the 3 HRRP conditions, we evaluated the average annual change in rates of care utilization across different care settings, based on ordinary least squares regression of rates of post-discharge care use—rates of readmissions, observation stays, ED visits, or care in either of 3 acute care settings—against calendar years. We used the non-parametric runs test to evaluate and confirm the linearity of calendar-year trends, which evaluates the randomness of distribution of data points around the regression line (see Online Supplement).(21) To better understand whether changes in patterns of care in a given post-acute care setting had progressed differently during the 30 days after discharge, we qualitatively evaluated care trends in these settings during post-discharge days 0-30 and 31-90 and whether there was an interaction between calendar year (2008-2016) and post-discharge period (day 0-30 versus 31-90) in analysis of covariance.

Next, we evaluated calendar-year and monthly trends in post-discharge 30-day mortality across the 4 patient groups, based on whether patients were readmitted, had an observation stay only, had an ED visit only, or had no care in any of these settings. We noted that these calendar-year trends were linear over the course of the study period based on visual assessment and using the runs test. We also confirmed normality of residuals and homoscedasticity of data, to ensure that assumptions of linear regression models were satisfied. We used an interaction term for post-discharge care group and calendar year in an analysis of covariance model to evaluate whether calendar-year trends in mortality were significantly different across the patient groups. We evaluated average annual changes in mortality rates across care settings, reported as the coefficient of change of the respective outcome in an ordinary least square regression with calendar year as a continuous independent variable. To evaluate the contribution of changes in mortality across individual care settings to the overall changes in mortality in the post-discharge 30-day period, we calculated the proportion of annual change in overall mortality rates that occurred across the 3 settings.

Next, to specifically address the possible associations with the announcement and implementation of HRRP with changing utilization across care settings, we constructed interrupted time series models, assessing how slope of the temporal trends in monthly rates of post-discharge care in the 3 acute care settings changed with the announcement and implementation of the HRRP. These analyses are consistent with the approach outlined in prior studies.(1, 9, 22)

Further, to evaluate whether systematic differences in patient characteristics were driving differences in outcomes, we evaluated differences in comorbidities and in-hospital events such as operative procedures and complications, as well as discharge disposition across these groups. We specifically evaluated whether comorbidities that are markers of debility, such as dementia, delirium, and encephalopathy, pressure ulceration, and protein calorie malnutrition, varied across groups. The comorbidities were ascertained from all claims across inpatient and outpatient care settings in the 1-year preceding the index hospitalization. The issues in the expansion of inpatient codes during this study period are mitigated with the use of comprehensive inpatient and outpatient codes. Finally, we assessed for markers for end-of-life care at discharge, including consultations for palliative care and discharge from hospital to hospice facilities.

All other analyses were performed using SAS, version 9.4 (Cary, NC), and Stata 14 (College Station, TX). The level of significance was set at 0.05. The study was reviewed by the Yale University Institutional Review Board and deemed exempt from informed consent due to the deidentified data. The study was reported in accordance with the STrengthening the Reporting of OBservational studies in Epidemiology (STROBE) recommendations, and its checklist is included in the online supplement.

### Patient involvement and Dissemination

No patients were involved in the development of the research question or the outcome measures, or in developing plans for the design and implementation of the study. The data are de-identified and, therefore, cannot be shared with the study participants directly. We will enlist patient and public participation in the dissemination of the results of the study.

## RESULTS

There were 3,772,924 hospitalizations for heart failure, 1,570,113 for AMI, and 3,131,162 for pneumonia during the study period. Of these, 22.5% patients with heart failure, 17.5% with AMI, and 17.2% with pneumonia were readmitted within 30 days of discharge. The overall rate of observation stays and ED visits were 1.7% and 6.4% for heart failure, 2.6% and 6.8% for AMI, and 1.4% and 6.3% for pneumonia, respectively. Cumulatively, a third of all hospitalizations—30.7% for heart failure, 26.9% for AMI, and 24.8% for pneumonia—received post-discharge care in any acute-care setting.

### Trends in Post-discharge Acute Care

The 30-day observed readmission rates decreased for all 3 conditions, from 23.5% in 2008 to 21.7% in 2016 for heart failure, 19.0% in 2008 to 15.9% in 2016 for AMI, and 17.6% in 2008 to 16.4% in 2016 for pneumonia (eFigure 1). Risk-adjusted rates followed a similar pattern, with a decrease from 24.5% to 21.0% for heart failure, 19.0% to 16.1% for AMI and 18.3% to 16.2% for pneumonia (eFigure 2). On average, risk-adjusted readmission rates decreased by 0.51% (95% CI, −0.66% to −0.35%) per year for heart failure, 0.44% (−0.56% to −0.32%) per year for AMI, and 0.33% (−0.43% to −0.23%) per year for pneumonia (Figure 1).

**Figure 1:**
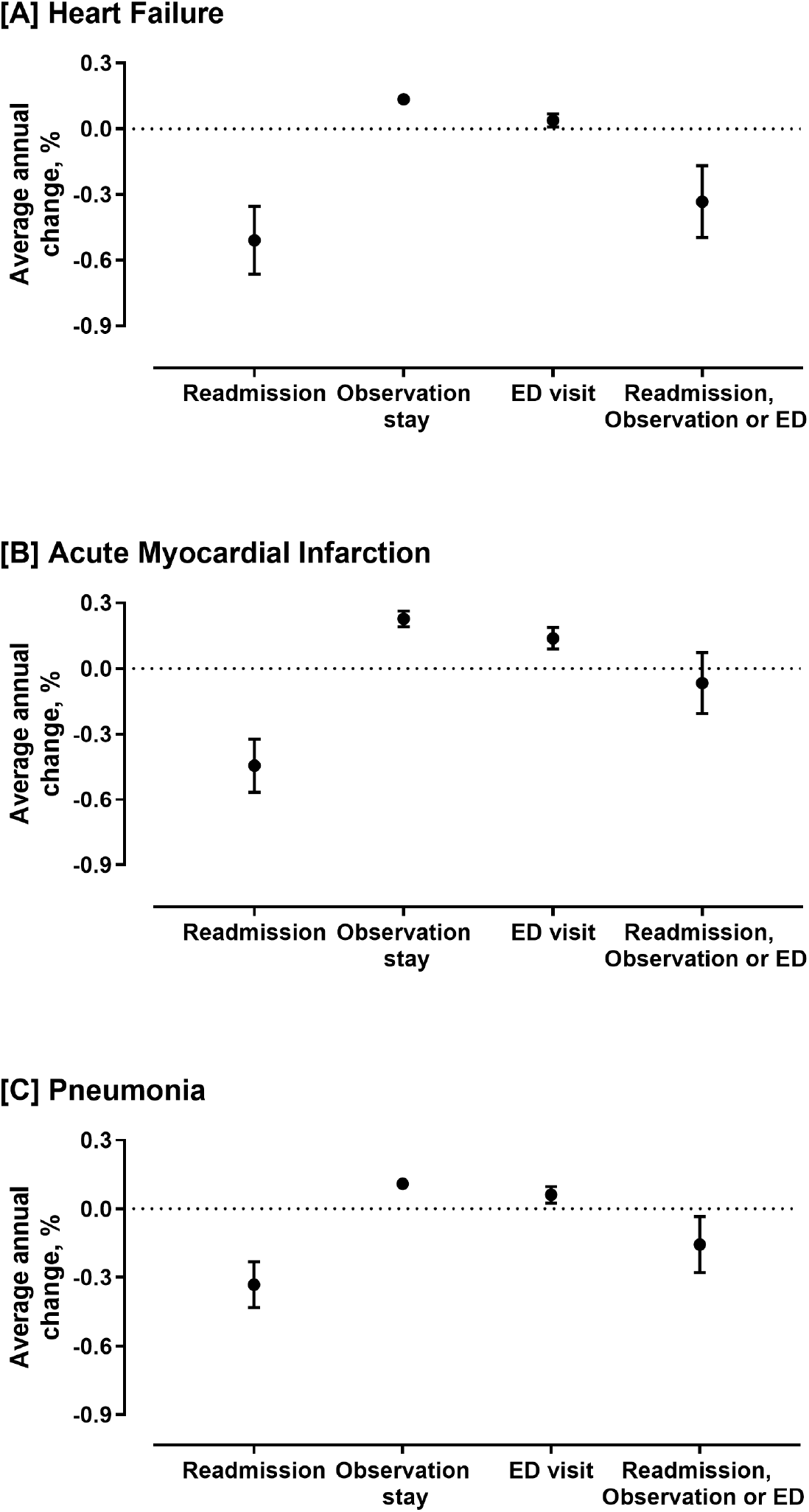
Average annual absolute percentage change in risk-adjusted post-discharge care use. Coefficient for annual change derived from ordinary least squares regression. Readmissions, observation stays, and emergency department (ED) visits are mutually exclusive, presented in order of hierarchy.

**Figure 2:**
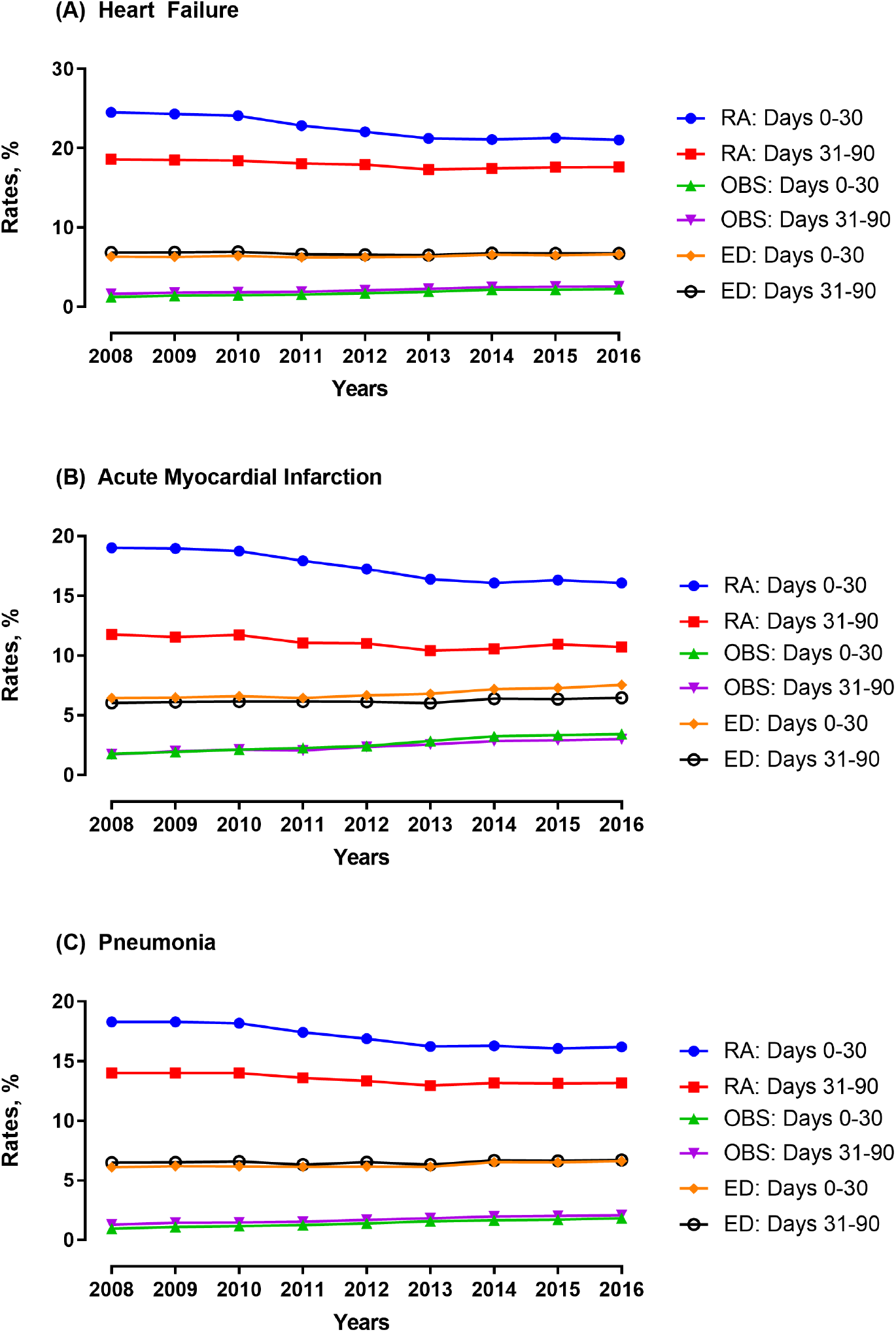
Calendar-year trends in risk-adjusted post-acute care utilization during post-discharge days 0-30 and 31-90. ED: emergency department, OBS: observation stay, RA: readmission

There was an increase in post-discharge 30-day observation stays and ED visits across the 3 conditions, with an average annual increase in risk-adjusted rates of observation stays of 0.13% (0.11% to 0.15%) for heart failure, 0.23% (0.19% to 0.26%) for AMI, and 0.11% (0.10% to 0.12%) for pneumonia among those not readmitted (P<0.001 for all). Similarly, there was a smaller but significant increase in rates of post-discharge 30-day ED visits for all 3 conditions, with an average annual increase of 0.04% (0.01% to 0.07%) for heart failure, 0.14% (0.09% to 0.19%) for AMI, and 0.06% (0.02% to 0.10%) for pneumonia (P<.001 for all) (Figure 1). While the combination of the increase in ED and observation stays was less than the decrease in readmission, the overall change in post-discharge acute care was not significant.

The increase in post-discharge observation stays and ED visits was also observed during days 31-90 post-discharge (Figure 2, eFigure 3). There was no significant difference between the increase in risk-adjusted rates of observation stays during post-discharge days 0-30 and 31-90, across calendar years for heart failure (P = 0.62) or pneumonia (P = 0.50). For AMI, the post-discharge observation stays increased in both periods, but at a higher rate during days 0-30 than days 31-90 (P for differences in slopes = 0.005). Similarly, ED visits increased during the first 30 days post-discharge as well as in the 31-90-day period, with a larger relative increase in the early post-discharge period across for heart failure and AMI (P for differences in slopes for HF, 0.02, and AMI, 0.003), but not for pneumonia (P = 0.13).

**Figure 3:**
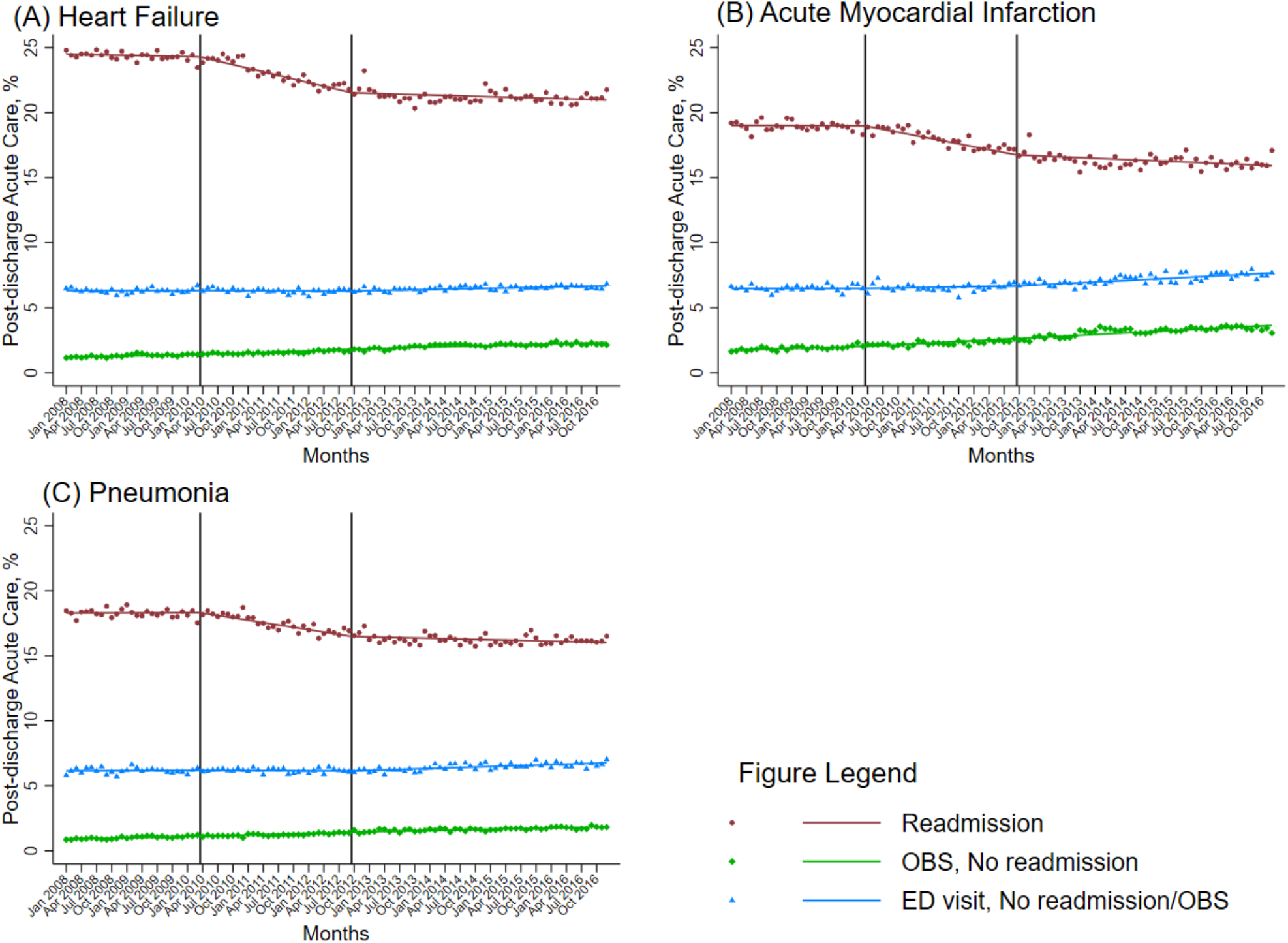
Changes in monthly rates of risk-adjusted post-acute care utilization after the announcement and implementation of the Hospital Readmissions Reduction Program (HRRP). Dots represent monthly risk-adjusted rates of post-acute care use in individual settings, with lines representing the slope of change in rates during each period relative to the HRRP. The interrupted time series models evaluated for changes in slopes at announcement and implementation of the HRRP, which are demarcated by vertical lines.

In interrupted time series models for both unadjusted and risk-adjusted assessments, monthly rates of post-discharge readmission rates decreased substantially at HRRP announcement across the 3 conditions (P for slope change, <0.001, Figure 3, eFigures 3-8, and eTable 4-9), and continued to decrease at a slower rate after HRRP’s implementation. The rates of observation stays were increasing before HRRP for all conditions. They accelerated after HRRP was announced, across both unadjusted and risk-adjustment assessments for heart failure. For AMI and pneumonia, while unadjusted observation rates did not demonstrate an inflection, conservative risk-adjustment assessments suggest an increase in observation stays that was approximately a fourth of the decrease in corresponding readmission rates (P for slope change <0.05, eTables 8 and 9). Across all 3 conditions, there were no inflection in rates of ED visits after HRRP announcement, but there was a significant increase after HRRP implementation that was approximately a fourth of the decrease in readmissions during this period (P for slope-change <0.05).

### Mortality Across Post-Discharge Care Groups

The overall post-discharge 30-day mortality was 8.7% for heart failure, 7.3% for AMI, and 8.4% for pneumonia. Post-discharge 30-day mortality was higher in those with readmissions (13.2% in heart failure, 12.7% for AMI, and 15.3% for pneumonia), compared with patients with observation stays (4.5% in heart failure, 2.7% for AMI, and 4.6% for pneumonia), or ED visits (9.7% in heart failure, 8.8% for AMI, and 7.8% for pneumonia), or no post-discharge acute care (7.2% in heart failure, 6.0% for AMI, and 6.9% for pneumonia).

Cumulatively, a third of post-discharge deaths occurred in individuals who were readmitted within the 30-day period (eFigure 9). Among those not readmitted, most deaths within the post-discharge 30-day period occurred in individuals who had neither observation stays nor ED visits, which represented more than 60% of the overall post-discharge 30-day mortality across the conditions. Those who were not readmitted but who received care in observation units or the ED represented less than 10% of all deaths in the post-discharge period (eFigure 9).

Over the study period, risk-adjusted post-discharge mortality increased from 8.4% to 8.8% for heart failure, decreased from 7.5% to 7.0% for AMI, and 8.3% to 8.1% pneumonia (Figure 4). The increase in post-discharge mortality for heart failure began before the announcement of the HRRP. The average risk-adjusted post-discharge 30-day mortality increased by 0.05% (95% CI, 0.02% to 0.08%) per year for heart failure and decreased by 0.06% (−0.09% to −0.04%) per year for AMI during 2008-2016 (Figure 5). Of the overall increase in mortality in heart failure, increase in mortality was concentrated among individuals who were neither readmitted nor received care in an observation unit or an ED (Figure 5). The rate of increase in mortality in this group without any acute care in either inpatient units, observation wards, or the ED was 0.08% per year (95% CI, 0.05% to 0.12%), rates of increase that exceeded the overall increase in post-discharge mortality in heart failure. In contrast, decrease in post-discharge mortality in AMI was concentrated among those that were readmitted. For pneumonia, the overall mortality rates did not change over the study period. Therefore, increase in post-discharge mortality were limited to patients with heart failure, who did not receive post-discharge acute care in of the 3 settings.

**Figure 4:**
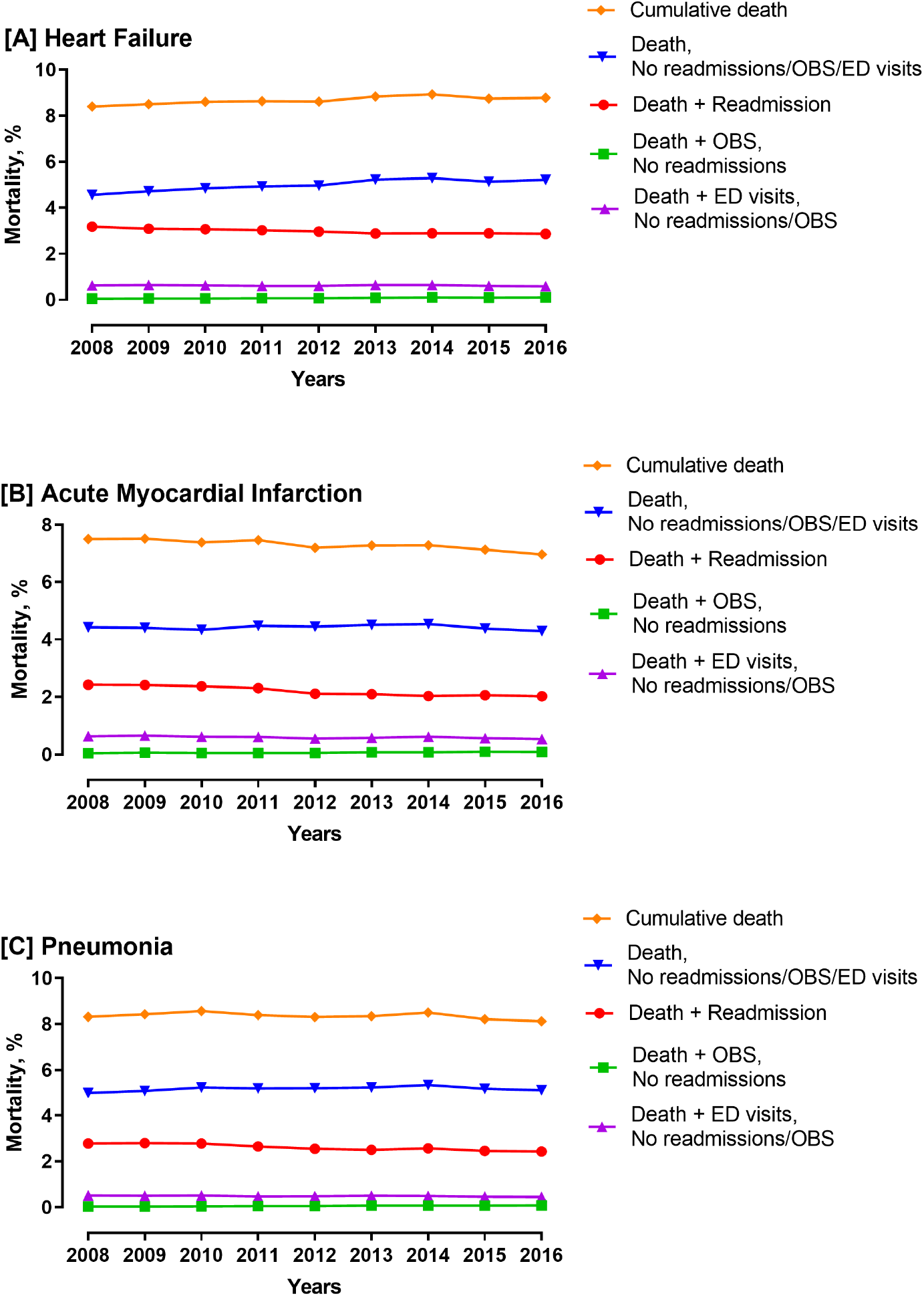
Risk-adjusted rates of death in the 30-day post-discharge period by post-discharge care use, 2008-2016. Points represent proportion of patients discharged alive who died in the post-discharge 30-day period, based on post-discharge acute care utilization (Abbreviations: ED: emergency department, OBS: observation stay).

**Figure 5:**
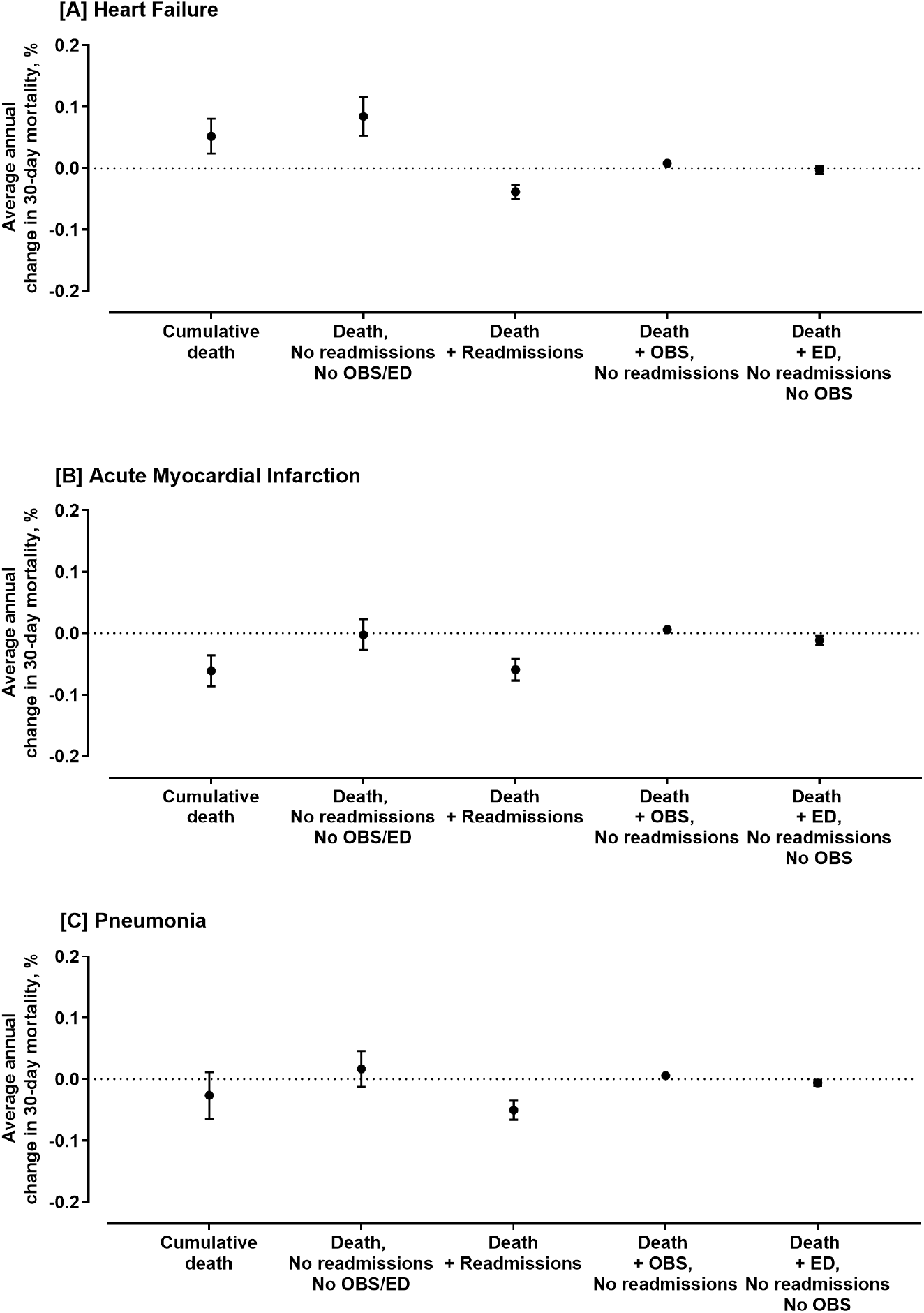
Average annual absolute percentage change in risk-adjusted 30-day post-discharge mortality by post-discharge care use. Coefficient for annual change derived from ordinary least squares regression. Readmissions, observation stays (OBS), and emergency department (ED) visits are mutually exclusive, presented in order of hierarchy.

There were no effects of HRRP announcement or implementation on temporal trends in mortality among patients receiving post-discharge acute care in observation units or the ED. Specifically, in interrupted time series models that evaluated for the effects of mortality, across all 3 conditions and for unadjusted as well as risk-adjusted assessments, there were no HRRP associated inflections in mortality in these settings at either the announcement of the HRRP or its implementation (Figure 6, eFigures 12-17, eTables 10-15). This suggests that the HRRP was not associated with an increase in mortality risk among patients receiving care in the observation units or the ED in the post-discharge period.

**Figure 6:**
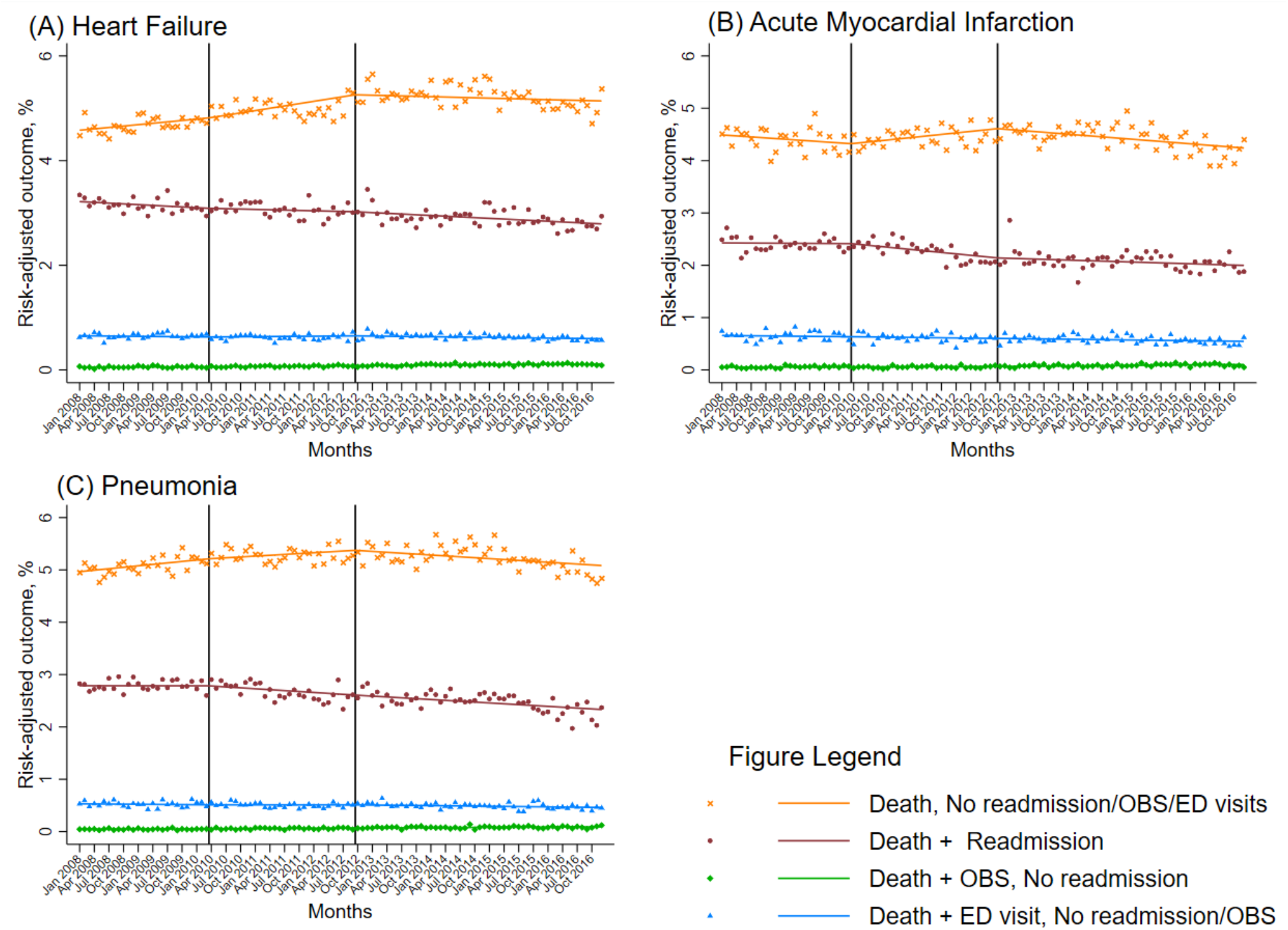
Changes in slopes of monthly rates of risk-adjusted post-discharge mortality across groups of post-discharge acute care utilization after the announcement and implementation of the Hospital Readmissions Reduction Program (HRRP). Dots represent monthly risk-adjusted rates of post-acute care use in individual settings, with lines representing the slope of change during each period relative to the HRRP. The interrupted time series models evaluated for changes in slopes at announcement and implementation of the HRRP, which are demarcated by vertical lines.

### Characteristics of Patients by Post-discharge Care Setting and Outcomes

There were differences in markers of frailty and advanced health care needs between groups based on whether they survived to post-discharge day 30 or died after seeking post-discharge care in various settings (Tables 1-3, eTables 16-18). Across all conditions, those who died had a higher prevalence of markers of debility including dementia, functional disability, delirium, chronic skin ulceration, and protein calorie malnutrition. Among the deceased, the prevalence rates of these conditions were substantially higher in the group of patients who died without having used any post-discharge acute care (Tables 1-3). Those that died also had a higher rate of markers of advanced disease such as cardiorespiratory failure, metastatic cancer, and acute leukemia; however, these markers did not differ between groups based on their post-discharge care use. Further, the group of patients without post-discharge acute care use had much higher rates of palliative care consultations and discharge to hospice. In the group of patients who died without using post-discharge acute care, 19.4% with heart failure, 21.0% with AMI, and 17.0% with pneumonia had a palliative care consultation before discharge. Notably, while 3.6% patients with heart failure, 3.2% patients with AMI and 3.1% patients were discharged to hospice following index hospitalization, in patients who died without using post-discharge acute care, 45.3% with heart failure, 49.1% with AMI, and 42.0% with pneumonia had been discharged to hospice after the index hospitalization.

There were also disease-specific temporal trends in hospice discharges. Across all patients, 3.6% with heart failure, 3.2% with AMI, and 3.2% with pneumonia were discharged to hospice, with an increase in hospice discharges in heart failure and pneumonia but not in AMI (Figure 7).

**Figure 7:**
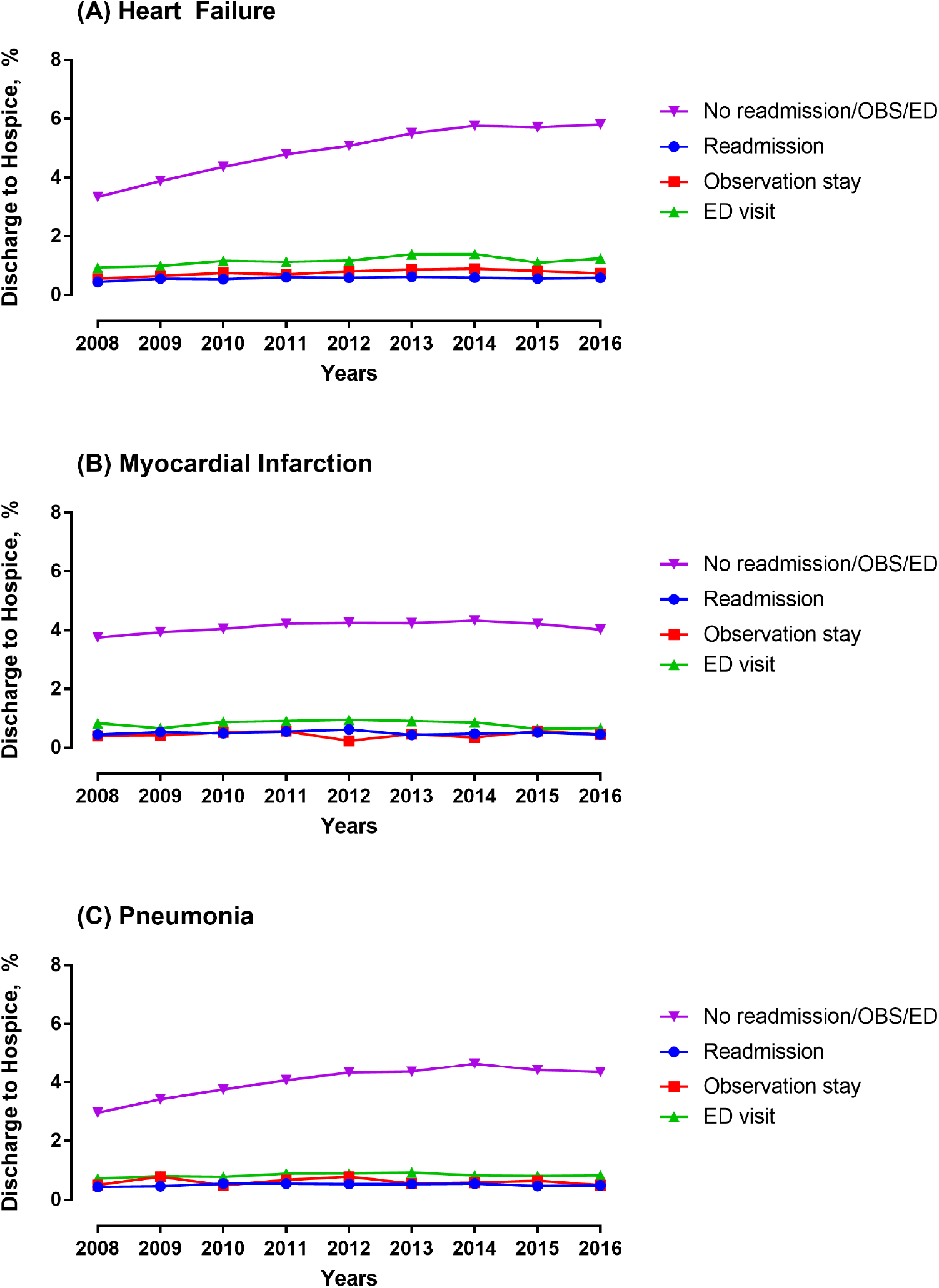
Trends in discharge to hospice. Discharge to hospice by calendar years across patient groups based on their post-discharge care utilization. ED: emergency department, OBS: observation stay

## DISCUSSION

Between 2008 and 2016, while the national readmission rate decreased significantly across all 3 conditions included in the HRRP, the use of the ED and observation units significantly increased. Although the readmission rate decline was greater than the rise in the other acute care settings, the overall post-discharge acute care change, representing the aggregate of readmission, observation stays and ED visits, did not significantly change. Patients receiving post-discharge care in the ED and observation units who were not readmitted had lower mortality than those who were readmitted. A majority of deaths in the 30-day post-discharge period occurred in patients who did not seek any post-discharge acute care, with fewer than 10% occurring in those with post-discharge care in observation units and the ED. During this period, which spanned before the HRRP through the announcement and implementation, only patients with heart failure had a significant increase in post-discharge mortality, which began before the HRRP announcement. The increase, however, was almost exclusively observed among those without post-discharge acute care and was not a result of deaths among those with ED or observation stays who were not admitted. Nearly half of all patients who died without any post-discharge acute care had been discharged to hospice from the index hospitalization.

### Trends in Post-discharge Observation Stays and ED visits

Observation stays and ED visits have increasingly been used in the post-discharge period.(1) We find that the rise in the care in these settings counterbalances improvements in readmissions during 2008-2016, expanding on a similar observation analysis using Medicare data from 2012-2015.(23) Collectively, the patterns suggest a transition in how patients receive post-discharge acute care. The MedPAC, a non-partisan organization that does analyses for the US Congress, evaluated the HRRP and similarly found reductions in readmissions with increases in ED and observation visits.(24) Notably, we found an increase in these care settings that was not restricted to the 30-day measure reporting period, and increased at a similar rate during days 31-90 post-discharge. The MedPAC, in addition, also found that observation stays and ED visits occurred among conditions that were not targeted in the HRRP.(24) Therefore, it is unclear whether the changing patterns of post-discharge acute care was part of a strategy to reduce readmission or whether they are related to other concurrent policy changes such as the wider implementation of criteria for inpatient hospitalizations that discouraged short hospital stays as inpatient hospitalizations.(3, 25, 26) Nevertheless, the overall rise in alternative post-discharge acute care in the 30-day period requires an assessment of whether patients experience improved with this transition, and if there are avenues to further improve their post-discharge care recovery.

### Outcomes in Observation Stays and ED visits

We found that patients that received care solely in observation units and ED had lower rates of mortality than those who were hospitalized, indicating that these settings were likely utilized for lower risk patients. Thus, the result does not support the concern that high-risk patients are denied access to the higher-level services they need.(5, 6) The changes in mortality—especially for patients with heart failure, who have experienced rising post-discharge mortality—largely occurred among individuals who had had no post-discharge acute-care utilization.

What remains unclear is whether these changes in post-discharge mortality, which is exclusively observed for patients with heart failure, represent a failure of appreciating that these individuals needed acute care or a group with expected deaths in the setting of comfort-centered, end-of-life care. We noted that this group of patients that had high rates of palliative care utilization and discharge to hospice facilities. However, these observations do not completely explain the elevated mortality risk in patients who do not utilize post-discharge acute care. Moreover, while suggested in a previous study,(5) patients with pneumonia have not had a consistent rise in post-discharge mortality in our more contemporary assessment.

### Comparison to Literature and Implications for Health Policy

The study strongly suggests that the HRRP did not lead to harm through inappropriate triage of high-risk patients to observation units and the ED, and therefore, represents evidence against calls to curtail the program due to this theoretical concern.(6) The concern was mainly raised by one study that ascribed a rising mortality in the period spanning the HRRP announcement and implementation to the effects of the program, also reporting that this increase was restricted to those not readmitted.(5) This study, like others,(9, 27-29) found that post-discharge mortality in these conditions was increasing nearly 3 years before HRRP announcement, but ascribed an excess increase to HRRP after modelling HRRP’s effects as changes across four 30-month periods discounting within period trends.(5) It also used inpatient data to define case mix and used a consistent number of codes after the expansion of coding slots on inpatient claims. The conclusions of the study contrasted with another that also found an increase in heart failure and pneumonia mortality during 2006-2014 but did not find inflections in the slopes of monthly rates of mortality in interrupted time series models, while specifically accounting for within-period trends.(9) Moreover, it used inpatient, outpatient and physician claims for risk adjustment, and used all claims data for covariate identification after code slots expanded, without any analytic differences emerging due to covariate selection strategies. Interestingly, a recent study by the authors whose initial study suggested a rise in mortality for heart failure and pneumonia in Medicare following HRRP implementation found no changes in mortality for these conditions after modeling them as monthly changes during 2012-2015,(23) despite including the same study population and the same data. The study also did not report mortality rates in observation units and the ED, despite a focus on the care in these settings.(23)

Other studies using national Medicare data have focused on temporal trends in subgroups of hospitals and patients,(30, 31) and are therefore, not directly comparable to the above studies focusing on overall trends. Finally, an analysis using 100 thousand hospitalizations among Medicare beneficiaries in a quality improvement registry drawn from 500 US hospitals, which captured an inconceivably low 2 heart failure hospitalizations on average per hospital per month, did not replicate the secular trends in heart failure readmissions or mortality nationally.(22, 32) It, therefore, cannot form the basis of generalizable conclusions about the effects of the program.(32) The reassuring findings from the current study complement hospital-level assessments, which have found that reduction in readmissions are on average associated with reductions in mortality.(33) There has also been no evidence to support the hypothesis of gaming of quality measures by delaying or deferring readmissions, as there are no discontinuities in readmission or mortality around day-30.(34)

Our study also explores other potential concurrent changes in patient care, such as the end-of-life care that may reflect in post-discharge outcomes. However, whether markers of debility or end-of-life care require consideration in measures cannot be directly inferred. Rates of these illnesses among those readmitted are low and because they are potentially related to the quality of in-hospital care processes, excluding them in the assessment of readmission measures may not be appropriate. Finally, the current mortality measures were designed with considerations for such care needs before the index hospitalization, and patients receiving hospice care are excluded from the measure population.(35, 36) These measures focus on post-admission 30-day mortality, which capture the entirety of the care experience of patients during hospitalization and in the post-discharge period. Hospitals, therefore, focus on this composite of mortality. The current discussion about HRRP has exclusively focused on post-discharge mortality, despite a notable concurrent trend for reduction in in-hospital deaths, which have reduced at a comparable rate as the rise in post-discharge deaths. An independent assessment by MedPAC also found that risk-adjusted cumulative in-hospital and post-discharge 30-day mortality have slightly decreased over the period spanning the introduction of the HRRP.(3, 37)

### Limitations

Our study has some limitations. We are unable to identify patterns of acute care during the index hospitalization that would be associated with a higher rate of post-discharge acute care in observation units and EDs, and whether these visits represented avenues for planned post-discharge follow-up care. Moreover, the proportion of these care encounters that were preventable remains poorly understood. Next, our assessment of discharge destination is exploratory, and does not track the patient’s location in the post-discharge period. Finally, we were unable to elucidate the cause of death among individuals who did not seek post-acute care but died after a hospitalization due to our use of deidentified data. However, this is unlikely to represent refusal of care, as there are specific laws against refusal of care of acutely ill patients requesting emergency care. Moreover, current quality measures do not disincentivize care in the ED or observation units, and there are no incentives for hospitals to refuse acute or emergent care. Also, our study focuses on all US hospitals, and not merely the hospital with the index hospitalization, and the other hospitals do not have a reason to refuse needed care. An investigation into these deaths remains an important unaddressed question.

## CONCLUSION

Among patients hospitalized with conditions targeted in the HRRP, a recent decrease in readmissions is balanced by an increase in post-discharge 30-day observation stays and ED visits, such that the overall post-discharge acute utilization has remained unchanged. However, there is no evidence for harm related to increasing care in alternative acute care settings, and patients who received care in observation units or EDs, and were not readmitted, had low rates of post-discharge 30-day mortality and were not affected by the announcement or the implementation of the HRRP.

## Data Availability

The data are proprietary to the Centers for Medicare and Medicaid Services and can be obtained from them directly. The statistical code is available from the corresponding author.

## Disclosures

All authors have completed the ICMJE uniform disclosure form at www.icmje.org/coi_disclosure.pdf. Dr. Krumholz is a recipient of a research grant, through Yale, from Medtronic and Johnson & Johnson (Janssen) to develop methods of clinical trial data sharing; was a recipient of a research grant, through Yale, from Medtronic and the Food and Drug Administration to develop methods for post-market surveillance of medical devices; was a recipient of a research agreement, through Yale, from the Shenzhen Center for Health Information for work to advance intelligent disease prevention and health promotion, and collaborates with the National Center for Cardiovascular Diseases in Beijing; received payment from the Arnold & Porter Law Firm for work related to the Sanofi clopidogrel litigation and from the Ben C. Martin Law Firm for work related to the Cook IVC filter litigation; chairs a cardiac scientific advisory board for UnitedHealth; is a participant/participant representative of the IBM Watson Health Life Sciences Board; is a member of the Advisory Board for Element Science, the Physician Advisory Board for Aetna, and the Advisory Board for Facebook; and is the founder of Hugo, a personal health information platform. Drs. Krumholz, Bernheim, and Lin, and Mr. Wang work under contract with the Centers for Medicare & Medicaid Services to develop and maintain performance measures that are publicly reported. The other authors report no potential conflicts of interest.

The study was conceived and conducted by the authors, and the Centers for Medicare & Medicaid Services played no role in its design and conduct; collection, management, analysis, and interpretation of the data; and preparation, review, or approval of the manuscript.

## Contributors

All authors were responsible for the study concept and design. RK, YW and HMK were responsible for the acquisition and analysis of data. All authors contributed to the interpretation of the data. RK drafted the manuscript. All authors critically revised the manuscript for important intellectual content and approved the final version. RK and HMK are guarantors. The corresponding author attests that all listed authors meet authorship criteria and that no others meeting the criteria have been omitted.

## Transparency

The lead author affirms that this manuscript is an honest, accurate, and transparent account of the study being reported; that no important aspects of the study have been omitted; and that any discrepancies from the study as planned (and, if relevant, registered) have been explained.

